# Genetic relations between leukocyte counts, type 1 diabetes, and coronary artery disease

**DOI:** 10.1101/2024.04.01.24305155

**Authors:** Jolade Adebekun, Ajay Nadig, Priscilla Saarah, Samira Asgari, Linda Kachuri, David A. Alagpulinsa

## Abstract

**Hypothesis/Aim:** Type 1 diabetes (T1D) is associated with excess coronary artery disease (CAD) risk even when known cardiovascular risk factors are accounted for. Genetic perturbation of hematopoiesis that alters leukocyte production is a novel independent modifier of CAD risk. We examined whether there are shared genetic determinants and causal relationships between leukocyte counts, T1D, and CAD.

**Methods:** Genome-wide association studies (GWAS) summary statistics were utilized to perform pairwise linkage disequilibrium score regression (LDSC) and heritability estimation from summary statistics (ρ-HESS) to respectively estimate the genome-wide and local genetic correlations, and two-sample Mendelian randomization (MR) to estimate the causal relationships between leukocyte counts (335 855 healthy subjects), T1D (18 942 cases, 501 638 controls), and CAD (122 733 cases, 424 528 controls).

**Results:** There was significant genome-wide genetic correlation (r_g_) between T1D and CAD (r_g_ = 0.088; P = 9.0e-03) and both diseases shared significant genome-wide genetic determinants with eosinophil count (r_g(T1D)_ = 0.093, P = 7.20e-03; r_g(CAD)_ = 0.092, P = 3.68e-06) and lymphocyte count (r_g(T1D)_ = −0.052, P = 2.80e-02; r_g(CAD)_ = 0.1761, P = 1.82e-15). Sixteen independent loci showed stringent Bonferroni significant local genetic correlations between leukocyte counts, T1D and/or CAD. *Cis*-genetic regulation of the expression levels of genes within loci that are shared between T1D and CAD were associated with both diseases as well as leukocyte counts, including *SH2B3*, *CTSH*, *MORF4L1*, *CTRB1*, *CTRB2*, *CFDP1*, and *IFIH1*. Genetically predicted lymphocyte, neutrophil, and eosinophil counts were associated with T1D and CAD (lymphocyte odds ratio (OR)_T1D_ = 0.667, P = 2.02e-19; OR_CAD_ =1.085, P = 2.67e-06; neutrophil OR_T1D_ = 0.82, P = 5.63e-05; OR_CAD_ = 1.17, P = 5.02e-14; and eosinophil OR_T1D_: 1.67, P = 4.45e-25; OR_CAD_: 1.07; P = 2.02e-03).

**Conclusions/Interpretations:** This study sheds light on shared genetic mechanisms that underlie T1D and CAD, which may contribute to their co-occurrence through regulation of gene expression and leukocyte counts. This study also identifies molecular targets for further investigation for disease prediction and potential drug discovery.

**Research in Context:** *What is already known about the subject?:*

- Genetic factors have been shown to contribute to the occurrence of coronary artery disease (CAD) in type 1 diabetes (T1D), but the mechanisms are unknown.
- Genetic perturbation of hematopoiesis that alters leukocyte production is associated with CAD risk and clinical observations have documented altered leukocyte counts in T1D. However, it is unknown whether altered leukocyte frequencies contribute to T1D and the co-occurrence of T1D and CAD or these reflect reverse causation.

*What is the key question?:*

- Do T1D and CAD share genetic determinants with leukocyte counts, and if so, are genetically predicted leukocyte counts associated with risk of T1D or CAD and their co-occurrence?

*What are the new findings?:*

- T1D and CAD share significant genetic architecture, and both diseases share significant genetic determinants with eosinophil and lymphocyte counts.
- Genetically predicted eosinophil, lymphocyte, and neutrophil counts are associated with risk of T1D and CAD.
- Genetic regulation of the expression levels of genes in shared loci between T1D and CAD are associated with both diseases and leukocyte counts.

*How might this impact on clinical practice in the foreseeable future?:*

- Genetic heritability for T1D is shared with CAD risk and leukocyte counts, and the counts of eosinophils, lymphocytes, and neutrophils are associated with both diseases.

## Introduction

Type 1 diabetes (T1D) results from autoimmune-mediated destruction of insulin-producing pancreatic beta cells. Epidemiological studies suggest that coronary artery disease (CAD) occurs in patients with T1D more than in any other disease [1, 2]. Even when hyperglycemia and other known cardiovascular risk factors are accounted for, patients with T1D still have an increased risk of CAD relative to the general population [3–5]. The residual risk factors and their mechanisms that contribute to CAD in T1D are poorly defined, which complicates disease prediction and treatment. Population-based genetic studies have shown that T1D is genetically linked to CAD independent of traditional cardiovascular risk factors [3]. However, how genetic factors contribute to the comorbidity between T1D and CAD remains undefined. Genetic perturbation of hematopoiesis that alters the production of leukocytes is a novel independent modifier of several clinical outcomes, including autoimmune diseases and CAD [6–8]. For instance, genetic mutations in hematopoietic stem and progenitor cells (HSPCs) that alter leukocyte production known as clonal hematopoiesis of indeterminate potential (CHIP) can independently increase the risk of CAD up to four-fold [8]. Interestingly, integrated genetic and epigenomic analyses have shown that T1D risk variants are enriched in open chromatin of multiple hematopoietic cell types, including HSPCs [9–11]. Analyses of atherosclerotic lesions in patients also revealed that CAD risk variants are enriched in open chromatin of multiple cell types, including multiple immune cell types [12, 13].

Genetic variants that impact multiple hematopoietic cell types normally affect progenitor cells (i.e., HSPCs) to direct the production of distinct lineages [14, 15]. Thus, the above observations suggest that genetic perturbation of the production of leukocytes may contribute to T1D and CAD and potentially, their co-occurrence. In line with this hypothesis, clinical observations have documented altered leukocyte counts that correlate with the progression of T1D [16–19] and CAD [20]. In particular, the counts of neutrophils, which constitute the bulk of circulating leukocytes and are short-lived, requiring continuous replenishment by hematopoiesis [21], are consistently diminished in genetically at-risk individuals for T1D and in those diagnosed with the disease [16–19]. However, it is unknown whether these altered leukocyte counts causally contributes to disease and the co-occurrence of T1D and CAD, or these altered leukocyte counts are simply a consequence of disease. Unlike observational studies, association studies that employ inherited genetic variants as instruments are not confounded by environmental or lifestyle and disease. In this study, we utilized large-scale genome-wide association studies (GWAS) summary statistics to perform statistical genetic analyses to assess the genetic correlations and the potential causal relationships between leukocyte counts, T1D, and CAD.

## RESEARCH DESIGN AND METHODS

### GWAS datasets and study design

The overall study design is illustrated in **Figure 1A**. For leukocyte counts, we used summary statistics of our previously published blood cell traits GWAS based on 335 855 UK Biobank (UKB) European participants who had no conditions or diseases that may influence complete blood counts [22]. GWAS summary statistics for T1D (18 942 cases; 501 638 controls) [9] and CAD (122 733 cases; 424 528 controls) [23], whose participants were also of European ancestry, were obtained from the GWAS catalog (accession numbers GCST90014023 and GCST005195, respectively).

**Figure 1.**
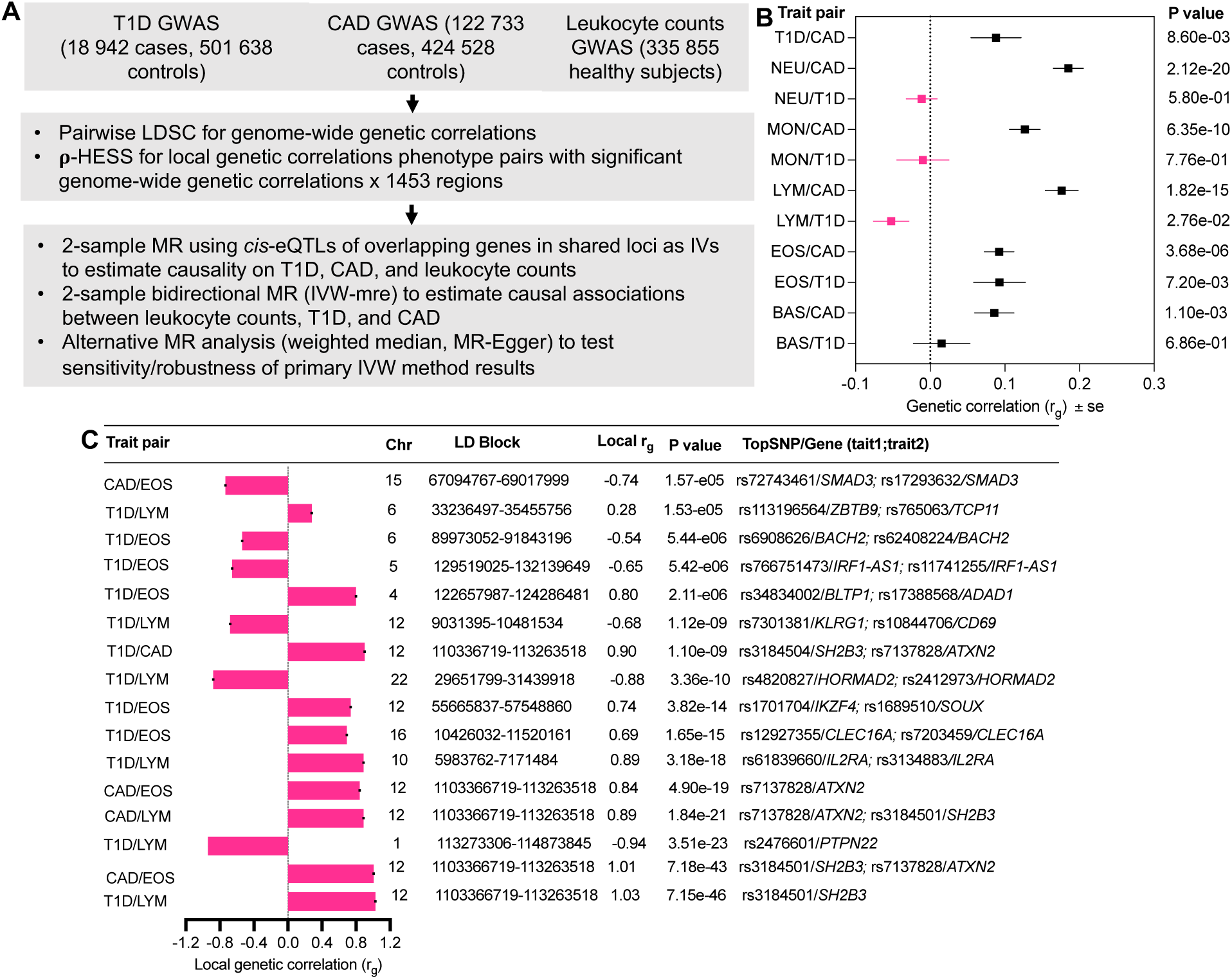
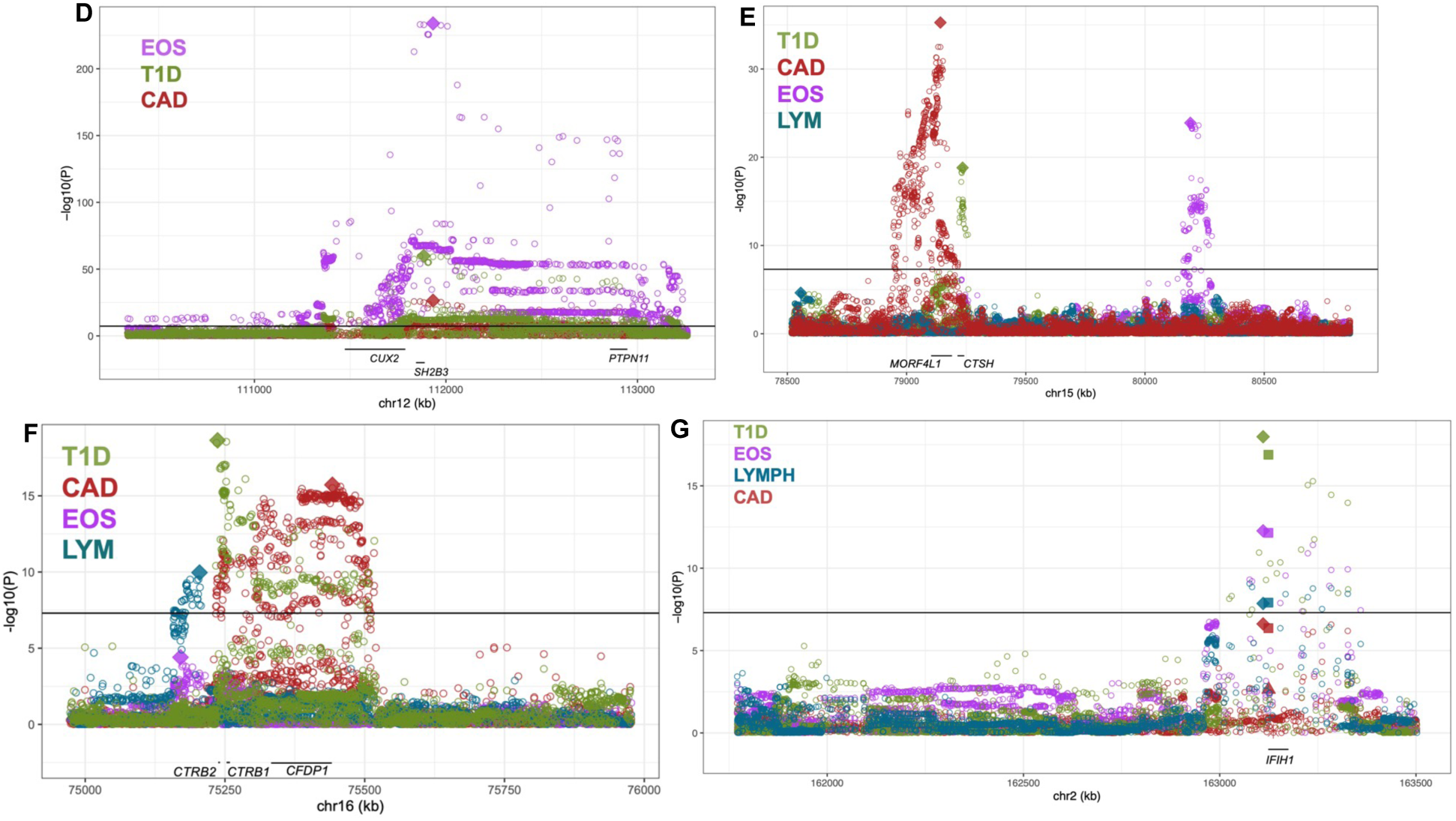
Genetic correlations between leukocyte counts, T1D, and CAD. (**A**) Schematic of study design. (**B**) Genome-wide genetic correlations (±se) between phenotypes estimated by LDSC. (**C)** Local genetic correlations between leukocyte counts, T1D, and/or CAD. **(D-G) Local genetic correlations between T1D, CAD, and leukocyte (eosinophil and lymphocyte) counts at genes of *apriori* interests.** Manhattan plots showing local genetic correlations between T1D, CAD, and leukocyte (EOS and LYM) counts at the **(D)** *SH2B3/CUX2/PTPN11* and (**C**) *IFIH1* **(D)** *MORF4L/CTSH* and **(E)** *CTRB2/CTRB1/CFDP1* loci. For each gene locus, local heritability and genetic correlation estimates are shown for each pair of traits for the LD-independent block containing the gene, with corresponding p values (−log10 P).

### Genetic correlation analyses

We conducted estimation of the genome-wide genetic correlations (r_g_) between leukocyte counts, T1D, and CAD using linkage disequilibrium score regression (LDSC). Analyses were performed using the HapMap3/1000 Genomes reference panel, restricting to variants with minor allele frequency (MAF) >0.01 outside of the major histocompatibility complex (MHC) region. Leukocyte counts included basophil count (BAS), eosinophil count (EOS), lymphocyte count (LYM), monocyte count (MON), and neutrophil count (NEU), yielding 11 phenotype pairs, i.e., T1D/CAD, T1D/BAS, T1D/EOS, T1D/LYM, T1D/MON, T1D/NEU, CAD/BAS, CAD/EOS, CAD/LYM, CAD/MON, and CAD/NEU.

In addition to genome-wide genetic correlation, locus-specific or local genetic correlation between phenotypes could provide further evidence of shared etiology. For instance, there may be some regions in the genome where genetic correlations between phenotypes differ in magnitude or sign from the genome-wide genetic correlation [24]. We performed local genetic correlation analyses using heritability estimation from summary statistics (ρ-HESS) [25], which uses a fixed-effect model to estimate heritability and genetic covariances within specific regions of the genome, while accounting for linkage disequilibrium (LD) and sample overlap [25]. Local genetic correlations between T1D, CAD, and leukocyte counts were estimated at 1453 LD-independent blocks across the genome, excluding the MHC region [26]. We conducted two types of analyses of the ρ-HESS results. We first undertook an agnostic genome-wide analysis approach to identify the blocks for each phenotype pair that had heritability and genetic covariance significantly different than 0 at a stringent Bonferroni threshold of p<0.05/1453 blocks (i.e., p<3.44e-05). In the second approach, we adopted a candidate gene approach, examining the local genetic correlations at blocks containing pre-selected genes based on previous studies that implicate these genes in either T1D, CAD or hematopoietic traits [6, 27, 28]. Local genetic correlations were only computed for candidate gene blocks with significant local heritability observed for both paired-phenotypes. After filtering for loci with non-significant local heritability, we performed 36 significance tests for local genetic covariance at candidate gene blocks and used a Bonferroni threshold of p < 0.05/36 (i.e. p < 1.34e-3).

### Two-sample Mendelian randomization (MR) analyses

All MR analyses were performed using the *TwoSampleMR* package in R.

#### Causality of shared gene loci between T1D and CAD

To infer the putative causality of loci that showed local genetic correlation between T1D and CAD, we used *cis*-expression quantitative trait loci (*cis*-eQTL) of the nominated/overlapping genes in the shared loci as instruments to perform the MR analyses because genetic variants underlying complex diseases such as T1D and CAD predominantly act as *cis*-regulatory elements [9, 29, 30]. We extracted *cis-*eQTLs (eSNPs) that significantly (p<1.0e-05; FDR<0.05) regulate the expression levels of the nominated genes from the Genotype-Tissue Expression (GTEx) consortium portal in disease-relevant tissue whenever available, including whole blood, pancreas, and EBV-transformed lymphoblastoid cell lines (LCLs). Using these eSNPs as instrumental variable (IVs), we performed two-sample MR by calculating the Wald ratios to estimate the effect of the expression levels of each gene on the risk of T1D, CAD, and leukocyte counts. The multiplicative random effects inverse variance weighted (IVW-mre) approach was performed to combine the Wald ratios of multiple *cis*-eQTLs to obtain an overall MR estimate for each gene.

#### Association between leukocyte counts, T1D, and CAD

To investigate the association between genetically inferred leukocyte counts and risk of T1D and CAD, we performed bi-directional MR analyses by first treating leukocyte counts as the exposure, followed by the reverse analyses, wherein leukocyte counts were outcomes and genetic predisposition to T1D and CAD were the exposures. IVs for each exposure were LD-independent SNPs that reached genome-wide significance (p< 5.0e-08) for the given phenotype. We have previously described the IVs for leukocyte counts [22]. The IVs for T1D and CAD were clumped with an LD threshold of r^2^<0.001 based on the European 1000 Genomes LD reference to obtain independent IVs. MR makes several strong and untestable assumptions. For instance, horizontal pleiotropy, which occurs when a genetic instrument influences the outcome directly or through any pathway other than through the exposure, is a major threat to valid inference in MR. Therefore, we used multiple analytic approaches that make different assumptions to assess the stability of the results. The IVW-mre method, which accounts for some heterogeneity due to the presence of potentially invalid instruments, was used as the primary method to estimate the causal relationships between phenotypes. Heterogeneity was assessed using the Cochran’s Q statistic, whereby a Cochran Q test p value <0.05 indicated the presence of significant heterogeneity. If there was evidence of significant heterogeneity, outliers contributing to heterogeneity were pruned using Radial IVW regression [31]. We then looked for consistency in the IVW-mre results based the weighted median test [32], which is less sensitive to the influence of outliers, and the MR-Egger test, which tests for the presence of directional pleiotropy and corrects the effect estimate if pleiotropy is detected.

Unless otherwise stated, the MR results are presented as odds ratio (OR) and 95% CI (CI) of the risk of the outcome per 1-standard deviation (1-SD) increase in genetically predicted levels of the exposure.

## RESULTS

### Genetic correlations between T1D and CAD, and leukocyte counts

Shown in **Figure 1B**, there was significant positive genome-wide genetic correlation (r_g_) between T1D and CAD (r_g_ = 0.088, P=0.009). Both T1D (r_g_ = 0.093, P = 7.20e-03) and CAD (r_g_ = 0.092, P = 3.68e-06) had significant positive genome-wide genetic correlations with eosinophil count. In contrast to eosinophil count, T1D had a significant negative genome-wide genetic correlation with lymphocyte count (r_g_ = −0.052, P = 2.80e-02), whereas CAD had a significant positive genome-wide genetic correlation with lymphocyte count (r_g_ = 0.1761, P = 1.82e-15; **Figure 1B**). T1D also showed a negative genome-wide genetic correlation with neutrophil count, albeit statistically insignificant (r_g_ = −0.0115, P = 5.080e-01; **Supplemental (Suppl) Table 1**). CAD had a significant positive genome-wide genetic correlation with all other leukocyte counts, including, basophil, monocyte, and neutrophil counts (r_g_ ≥ 0.0858, P<0.0011; **Figure 1B** and **Suppl Table 1**).

**Table 1:** Associations between genetically determined expression levels of genes within loci that are shared between T1D and CAD and risk of both diseases and leukocyte counts.

| Gene | Tissue | No. SNPs | T1D OR(95%CI);<br>P value | CAD OR(95%CI);<br>P value | BAS OR(95%CI);<br>P value | EOS OR(95%CI);<br>P value | LYM OR(95%CI);<br>P value | MON OR(95%CI);<br>P value | NEU OR(95%CI);<br>P value |
| --- | --- | --- | --- | --- | --- | --- | --- | --- | --- |
| *SH2B3* | Blood | 10 | 0.30(0.25-0.37);<br>3.50e-31 | 0.83(0.78-0.88);<br>1.24e-10 | 0.93(0.90-0.96);<br>5.18e-07 | 0.65(0.59-0.72);<br>2.36e-16 | 0.66(0.58-0.75);<br>8.62e-11 | 0.90(0.84-0.97);<br>3.37e-03 | 0.89(0.85-0.94);<br>4.00e-05 |
| *SH2B3* | Pancreas | 291 | 0.64(0.63-0.65);<br><2.23e-308 | 0.89(0.89-0.89);<br><2.23e-308 | 0.96(0.96-0.96);<br>2<.23e-308 | 0.84(0.84-0.85);<br><2.23e-308 | 0.86(0.86-0.86);<br><2.23e-308 | 0.92(0.92-0.92);<br><2.23e-308 | 0.94(0.94-0.94);<br><2.23e-308 |
| *CTSH* | Blood | 287 | 1.45[1.43-1.47];<br>2.23e-308 | 0.81(0.80-0.82);<br>4.10e-221 | 1.00(0.99-1.00);<br>1.35e-10 | 1.00(1.00-1.00);<br>6.68e-01 | 0.99(0.99-0.99);<br>1.41e-14 | 0.99(0.99-0.99);<br>7.82e-21 | 0.98(0.98-0.98);<br>2.60e-130 |
| *CTSH* | LCL | 61 | 1.18(1.17-1.19);<br>9.40e-255 | 0.97(0.96-0.97);<br>6.90e-156 | 1.00(1.00-1.00);<br>3.15e-08 | 1.00(1.00-1.00);<br>8.86e-44 | 1.00(1.00-1.00);<br>1.96e-21 | 1.00(1.00-1.00);<br>1.11e-03 | 0.99(0.99-0.99);<br>3.80e-108 |
| *CTSH* | Pancreas | 64 | 1.27(1.26-1.29);<br>6.30e-221 | 0.92(0.92-0.93);<br>8.10e-122 | 1.00(1.00-1.00);<br>3.67e-01 | 1.00(0.99-1.00);<br>9.74e-04 | 1.00(1.00-1.00);<br>2.45e-01 | 1.00(1.00-1.00);<br>5.95e-09 | 0.99(0.99-0.99);<br>2.46e-26 |
| *MORF4L1* | Blood | 82 | 1.86(1.81-1.90);<br><2.23e-308 | 0.54(0.53-0.55);<br><2.23e-308 | 0.96(0.96-0.97);<br>1.70e-104 | 1.02(1.01-1.03);<br>4.59e-12 | 1.01(1.01-1.01);<br>5.93e-17 | 0.93(0.93-0.94);<br><2.00e-228 | 0.94(0.94-0.94);<br><2.23e-308 |
| *MORF4L1* | LCL | 35 | 0.96(0.95-0.97);<br>2.50e-10 | 1.22(1.22-1.23);<br><2.23e-308 | 1.01(1.01-1.01);<br>3.69e-13 | 1.00(1.00-1.00);<br>1.58e-01 | 1.02(1.02-1.02);<br>5.15e-86 | 1.03(1.02-1.03);<br>2.84e-91 | 1.02(1.01-1.02);<br>3.81e-25 |
| *MORF4L1* | Pancreas | 198 | 1.20(1.19-1.21);<br>3.10e-301 | 0.87(0.86-0.88);<br>9.60e-211 | 0.99(0.99-0.99);<br>4.55e-79 | 1.01(1.01-1.02);<br>6.55e-64 | 1.01(1.00-1.01);<br>2.06e-47 | 0.98(0.98-0.98);<br>2.40e-162 | 0.98(0.98-0.98);<br>2.80e-270 |
| *CFDP1* | Blood | 573 | 1.23(1.23-1.24);<br><2.23e-308 | 0.80(0.79-0.80);<br><2.23e-308 | 0.99(0.99-0.99);<br>1.70e-146 | 1.00(1.00-1.00);<br>1.96e-28 | 0.97(0.97-0.97);<br><2.23e-308 | 0.98(0.98-0.98);<br><2.23e-308 | 0.98(0.97-0.98);<br><2.23e-308 |
| *CFDP1* | LCL | 25 | 0.82(0.81-0.83);<br>2.23e-308 | 1.14(1.14-1.14);<br>2.23e-308 | 1.01(1.00-1.01);<br>4.44e-48 | 1.00(1.00-1.00);<br>3.69e-31 | 1.01(1.01-1.01);<br>2.23e-308 | 1.01(1.01-1.01);<br>3.62e-33 | 1.01(1.01-1.01);<br>4.86e-53 |
| *CFDP1* | pancreas | 43 | 1.38(1.36-1.40);<br><2.23e-308 | 0.81(0.80-0.81);<br><2.23e-308 | 0.99(0.99-0.99);<br>1.03e-18 | 1.00(1.00-1.00);<br>5.90e-18 | 0.98(0.98-0.99);<br>4.33e-69 | 0.99(0.99-1.00);<br>2.73e-11 | 0.99(0.99-0.99);<br>2.70e-20 |
| *CTRB1* | Pancreas | 79 | 0.88(0.87-0.89);<br>4.30e-134 | 0.92(0.91-0.92);<br>6.43e-94 | 1.01(1.01-1.01);<br>1.95e-25 | 0.98(0.97-0.98);<br>5.53e-80 | 0.99(0.99-0.99);<br>1.16e-49 | 0.99(0.99-0.99);<br>1.99e-62 | 1.01(1.01-1.01);<br>2.13e-31 |
| *CTRB2* | Pancreas | 172 | 1.04(1.04-1.05);<br>4.19e-58 | 1.03(1.03-1.03);<br>1.89e-70 | 1.00(1.00-1.00);<br>2.53e-22 | 1.01(1.01-1.01);<br>2.01e-91 | 1.00(1.00-1.00);<br>4.70e-13 | 1.01(1.01-1.01);<br>8.83e-25 | 1.00(0.99-1.00);<br>7.39e-40 |
| *IFIH1* | Blood | 11 | 1.12(1.11-1.14);<br>3.28e-42 | 1.24(1.04-1.49);<br>1.67e-02 | 0.97(0.97-0.98);<br>5.24e-19 | 1.02(1.01-1.02);<br>5.50e-07 | 1.03(1.03-1.04);<br>8.22e-31 | 0.98(0.97-0.98);<br>2.44e-53 | 0.98(0.98-0.99);<br>1.42e-14 |
| *IFIH1* | Pancreas | 18 | 0.99(0.99-1.00);<br>3.45e-02 | 1.02(1.02-1.02);<br>9.11e-73 | 1.00(1.00-1.00);<br>1.46e-01 | 1.02(1.02-1.02);<br>4.90e-152 | 1.03(1.03-1.03);<br>2.23e-308 | 1.01(1.01-1.01);<br>4.42e-89 | 1.02(1.01-1.02);<br>9.97e-83 |

Only eosinophil and lymphocyte counts were included in the local genetic correlation analyses because only these showed significant genome-wide genetic correlations with both T1D and CAD. The agnostic scan across the 1453 LD-independent blocks identified 16 genomic loci with Bonferroni significant (p<3.44e-05) local genetic correlation and local SNP heritability between phenotype pairs, i.e., between eosinophil and/or lymphocyte counts and T1D and /or CAD (**Figure 1C; Suppl Data 1**). There were an additional 55 genomic loci that showed nominally significant (p<0.05) local genetic correlations and local SNP heritability between phenotype pairs (**Suppl Data 1**).

Of the 16 significantly locally correlated loci, five were at an LD block on chromosome 12, of which Chr12:110336719-113263518 that harbors rs3184504 and rs7137828, respectively overlapping *SH2B3* and *ATXN2*, showed significant local genetic correlation across all four phenotypes, i.e., eosinophil count, lymphocyte count, T1D, and CAD (**Figure 1D; Suppl Data 1**). In addition to the *SH2B3/ATXN2* locus, two other genomic loci showed local genetic correlation between T1D and CAD at nominal significance, including Chr15:78516053-80860978 that harbors rs345593439 and rs7173743 that overlap *CTSH* and *MORF4L1* respectively (local r_g(T1D-CAD)_ = −0.4709, P_ρ-HESS_ = 1.87e-02; **Figure 1E; Suppl Data 1**), and Chr16:74971503-75977954, which harbors rs55993634 and rs8046696 that respectively overlap *CTRB2* and *CFDP1* (local r_g(T1D-CAD)_ = −0.6556, P_ρ-HESS_ = 1.93e-02; **Figure 1F; Suppl Data 1**). Of the 16 significantly shared loci (**Figure 1C**), Chr15:67094767-69017999 (rs72743461-*SMAD3* and rs17293632-*SMAD3*) was shared between CAD and eosinophil count only, while the rest of the 16 loci showed local genetic correlations between T1D and leukocyte counts (eosinophil and lymphocyte counts) only. These include *IKZF4*/*SUOX* (12:55665837-57548860) *CLEC16A* (Chr16:10426032-11520161), *HORMAD2* (22:29651799-31439918), *IRF1-AS1*(5:129519025-132139649), *BACH2* (6:89973052-91843196), *PTPN22* (1:113273306-114873845), *BLTP1/ADAD1/IL21*(4:122657987-124286481), *ZBTB9*/*TCP11*/*BAK1* (6:33236497-35455756), *IL2RA* (10:5983762-7171484).

The ρ-HESS analysis at 23 *apriori* selected gene loci corroborated the strong local genetic overlap and covariance across T1D, CAD, and eosinophil and lymphocyte counts at the LD-independent block on chromosome 12 that contains genes including *SH2B3*, *PTPN11*, and *CUX2* (local r_g_>0.8; P<1.0e-08; **Figure 1D; Suppl Data 1**). The *IFIH1* locus on Chr 2 also showed local genetic correlation across eosinophil count, lymphocyte count, T1D and CAD at nominal significance (**Figure 1G; Suppl Data 1**), which was also identified by the agnostic ρ-HESS analysis, i.e., Chr2: 161769733-163503551 harboring rs2111485-*IFIH1* and rs2075302-*IFIH1* (local r_g(T1D-EOS)_ = −0.5328, P_ρ-HESS_ = 3.00e-03; local r_g(T1D-LYM)_ = −0.4626, P_ρ-HESS_ = 6.40e-03; **Suppl Data 1**). However, while the local genetic correlation between T1D and CAD was statistically significant at the *IFIH1* locus (P_ρ-HESS_ = 1.67e-02), the local heritability for CAD did not reach the significance threshold (**Suppl Data 1**). The *CTRB1* locus on Chr16:75255860 also showed nominally significant local genetic correlation between T1D and CAD (local r_g_ = −0.6456, P_ρ-HESS_ = 1.93e-02). Other loci that showed local genetic correlation with either CAD and leukocyte counts or T1D and leukocyte counts include *TYK2-*Chr19:10476280.5 (local r_g(T1D-LYM)_ = −0.5982, P_ρ-HESS_ = 8.42e-05), *JAK1*-Chr1:65365549.5 local r_g(T1D-EOS)_ = 0.7810, P_ρ-HESS_ = 5.25e-05), and *IKZF3*-Chr17:37970819.5 (local r_g(T1D-LYM)_ = −0.7399, P_ρ-HESS_ = 2.73e-03). Of note, these ρ-HESS analyses results do not include several cases where significant local genetic covariance was observed in the setting of non-significant local heritability, as we elected conservatively not to interpret such cases (point estimates are included in **Suppl Data 1)**.

### Genetically determined expression of genes in loci shared between T1D and CAD is associated with risk of both diseases and leukocyte counts

The results for the effects of *cis-*eQTL-mediated expression of shared genes between T1D and CAD are summarized in **Table 2**. Overall, genetically determined expression levels of genes in shared loci between T1D and CAD were associated with risk of both diseases, either in the same or opposing directions, as well as altered leukocyte counts. *Cis*-eQTL-mediated upregulation of blood expression levels of *SH2B3*, whose locus positively overlapped across T1D, CAD, and leukocyte counts (EOS and LYM), were associated with reduced risk of both T1D (OR = 0.30, CI = 0.25-0.37, P = 3.50e-31) and CAD (OR = 0.80, CI = 0.72-0.88, P = 5.69e-06), and altered counts of all leukocyte types except monocytes (**Table 2**). The results for genetically determined increased expression levels of *SH2B3* in pancreas were similar to that for blood (**Table 2**). Genetically determined increased blood and pancreas expression levels of *CTSH*, *MORF4L1*, and *CFDP1*, whose loci showed negative local genetic correlation between T1D and CAD, also had opposing causal effects on the two diseases. Thus, while genetically determined increased blood and pancreas expression levels of *CTSH* were associated with increased T1D risk (OR_blood_ = 1.45, CI = 1.43-1.47, P <2.23e-308; OR_pancreas_ = 1.27, CI = 1.26-1.29, P = 6.30e-221), the risk of CAD was reduced (OR_blood_ = 0.81, CI = 0.80-0.82, P = 4.10e-221; OR_pancreas_ = 0.92, CI = 0.92-0.93, P = 8.10e-122). Likewise, genetic upregulation of blood and pancreas expression levels of *CFDP1* were associated with increased risk of T1D (OR_blood_ = 1.23, CI = 1.23-1.24, P <2.23e-308; OR_pancreas_ = 1.38, CI = 1.36-1.40, P <2.23e-308) but reduced CAD risk (OR_blood_ = 0.80, CI = 0.79-0.80, P <2.23e-308; OR_pancreas_ = 0.81, CI = 0.80-0.81, P <2.23e-308). Similarly, while *cis-*genetic upregulation of the expression levels of *MORF4L1* in blood and pancreas were associated with increased T1D risk (OR_blood_ = 1.86, CI = 1.81-1.90, P <2.23e-308; OR_pancreas_ = 1.20, CI = 1.19-1.21, P = 3.10e-301), the risk of CAD was reduced (OR_blood_ = 0.54, CI = 0.53-0.55, P <2.23e-308; OR_pancreas_ = 0.87, CI = 0.86-0.88, P = 9.60e-211). Unlike *CTSH* where genetic upregulation in blood, pancreas, and LCLs had the same direction of effects on T1D and CAD, upregulation of *CFDP1* and *MORF4L1* in LCLs had an opposing direction of effect compared to that in blood and pancreas, wherein genetically determined higher levels of *CFDP1* and *MORF4L1* in LCLs were associated with reduced T1D risk (OR_CFDP1_ = 0.96, CI = 0.95-0.97, P = 2.50e-10; OR_MORF4L1_ = 0.96, CI = 0.95-0.97, P = 2.50e-10) and increased CAD risk (OR_CFDP1_ = 1.15, CI = 1.11-1.20, P = 2.64e-15; OR_MORF4L1_ =1.22 CI = 1.22-1.23, P <2.23e-308). Whereas *cis-*genetic upregulation of pancreas expression levels of *CTRB1* were associated with reduced risk of both T1D (OR = 0.83, CI = 0.80-0.86, P = 4.59e-23) and CAD (OR = 0.92, CI = 0.91-0.93, P = 1.53e-70), upregulation of pancreas expression levels of *CTRB2* were associated with increased risk of both T1D (OR = 1.07, CI = 1.06-1.08, P = 5.87e-34) and CAD (OR = 1.02, CI = 1.02-1.03, P = 2.04e-25). We did not find *cis*-eQTLs for *CTRB1* and *CTRB2* in blood or LCLs from GTEx. Genetically determined expression levels of *CTSH, MORF4L, CFDP1, CTRB1, CTRB2* were also associated with leukocyte counts, except blood expression of *CTSH* that did not associate with eosinophil count; pancreas expression of *CTSH* that did not affect basophil and lymphocyte counts; and pancreas expression of *CTRB2*, which did not affect lymphocyte count.

**Table 2:** Association between genetically predicted leukocyte counts and risk of T1D and CAD.

| Exposure | No. SNPs | Outcome | MR test | OR | 95%CI | P value | Egger_intercept | P value |
| --- | --- | --- | --- | --- | --- | --- | --- | --- |
| BAS | 104 | T1D | IVW | 0.70 | 0.58-0.83 | 5.57e-05 | 3.17E-03 | 4.09E-01 |
|  | 104 |  | Weighted Median | 0.65 | 0.47-0.89 | 8.37e-03 |  |  |
|  | 104 |  | MR-Egger | 0.60 | 0.40-0.89 | 1.36e-02 |  |  |
| EOS | 231 | T1D | IVW | 1.67 | 1.52-1.85 | 5.45e-25 | -2.45E-03 | 4.28E-01 |
|  | 231 |  | Weighted Median | 1.50 | 1.29-1.75 | 1.88e-07 |  |  |
|  | 231 |  | MR-Egger | 1.83 | 1.44-2.31 | 1.10e-06 |  |  |
| LYM | 273 | T1D | IVW | 0.67 | 0.61-0.73 | 2.02e-19 | 7.59E-04 | 7.74E-01 |
|  | 273 |  | Weighted Median | 0.70 | 0.61-0.81 | 1.00e-06 |  |  |
|  | 273 |  | MR-Egger | 0.65 | 0.53-0.79 | 2.55e-05 |  |  |
| MON | 390 | T1D | IVW | 1.01 | 1.00-1.03 | 8.99e-03 | -1.17E-03 | 2.56E-01 |
|  | 390 |  | Weighted Median | 1.01 | 1.00-1.03 | 1.62e-02 |  |  |
|  | 390 |  | MR-Egger | 1.02 | 1.00-1.03 | 6.25e-03 |  |  |
| NEU | 217 | T1D | IVW | 0.82 | 0.75-0.90 | 5.63e-05 | -4.80E-03 | 7.68E-02 |
|  | 217 |  | Weighted Median | 0.84 | 0.72-0.98 | 2.65e-02 |  |  |
|  | 217 |  | MR-Egger | 0.97 | 0.79-1.18 | 7.32e-01 |  |  |
| BAS | 112 | CAD | IVW | 1.23 | 1.13-1.35 | 2.47e-06 | -7.04E-04 | 7.09E-01 |
|  | 112 |  | Weighted Median | 1.21 | 1.05-1.38 | 6.50e-03 |  |  |
|  | 112 |  | MR-Egger | 1.27 | 1.05-1.54 | 1.42e-02 |  |  |
| EOS | 286 | CAD | IVW | 1.07 | 1.03-1.11 | 2.03e-04 | -5.97E-04 | 6.03E-01 |
|  | 286 |  | Weighted Median | 1.02 | 0.96-1.09 | 4.70e-01 |  |  |
|  | 286 |  | MR-Egger | 1.09 | 1.01-1.18 | 3.20e-02 |  |  |
| LYM | 323 | CAD | IVW | 1.09 | 1.05-1.12 | 2.67e-06 | 4.98E-04 | 6.51E-01 |
|  | 323 |  | Weighted Median | 1.07 | 1.01-1.13 | 2.40e-02 |  |  |
|  | 323 |  | MR-Egger | 1.07 | 0.99-1.16 | 1.07e-01 |  |  |
| MON | 404 | CAD | IVW | 1.00 | 0.99-1.00 | 5.26e-01 | 6.86E-04 | 1.38E-01 |
|  | 404 |  | Weighted Median | 1.00 | 0.99-1.00 | 4.08e-01 |  |  |
|  | 404 |  | MR-Egger | 1.00 | 0.99-1.00 | 3.45e-01 |  |  |
| NEU | 241 | CAD | IVW | 1.17 | 1.12-1.21 | 5.02e-14 | -7.15E-04 | 5.44E-01 |
|  | 241 |  | Weighted Median | 1.14 | 1.06-1.23 | 3.15e-04 |  |  |
|  | 241 |  | MR-Egger | 1.19 | 1.10-1.29 | 3.38e-05 |  |  |

Genetically determined increased blood expression levels of *IFIH1* were associated with increased risk of both T1D (OR = 1.12, CI = 1.11-1.14, P = 3.28e-28) and CAD (OR = 1.24, CI = 1.04-1.49, P = 1.67e-02), as well as altered leukocyte counts. However, while genetically predicted transcript levels of *IFIH1* in the pancreas were associated with risk of CAD (OR = 1.02, CI = 1.02-1.02, P = 9.11e-73), the risk of T1D was insignificant (OR = 0.99, CI = 3.45e-02). Given that *IFIH1* is an established T1D susceptibility gene, the finding that *cis*-genetic upregulation of *IFIH1* expression levels in blood increases both T1D and CAD risk led us to investigate whether genetic variants in *IFIH1* that are associated with T1D risk also influence CAD risk. Using T1D GWAS significant (p<5.0e-08) SNPs that reside within the *IFIH1* gene and impact transcript and/or protein function [9, 11, 33, 34] as genetic instruments, we found that *IFIH1* variants that increase T1D risk also increase CAD risk (OR = 1.16, CI = 1.11-1.21, P = 5.83e-10; **Figure 2**) and alter leukocyte counts, including decreased counts of basophils (OR = 0.96, CI = 0.95-0.98, P = 5.97e-06), eosinophils (OR = 0.92, CI = 0.89-0.96, P = 2.64e-05), lymphocytes (OR = 0.89, CI = 0.87-0.91, P = 5.98e-31), and neutrophils (OR = 0.95, CI = 0.92-0.97, P = 6.68e-05), except monocytes (OR = 1.00, CI = 0.99-1.01, P = 8.40e-01).

**Figure 2:**
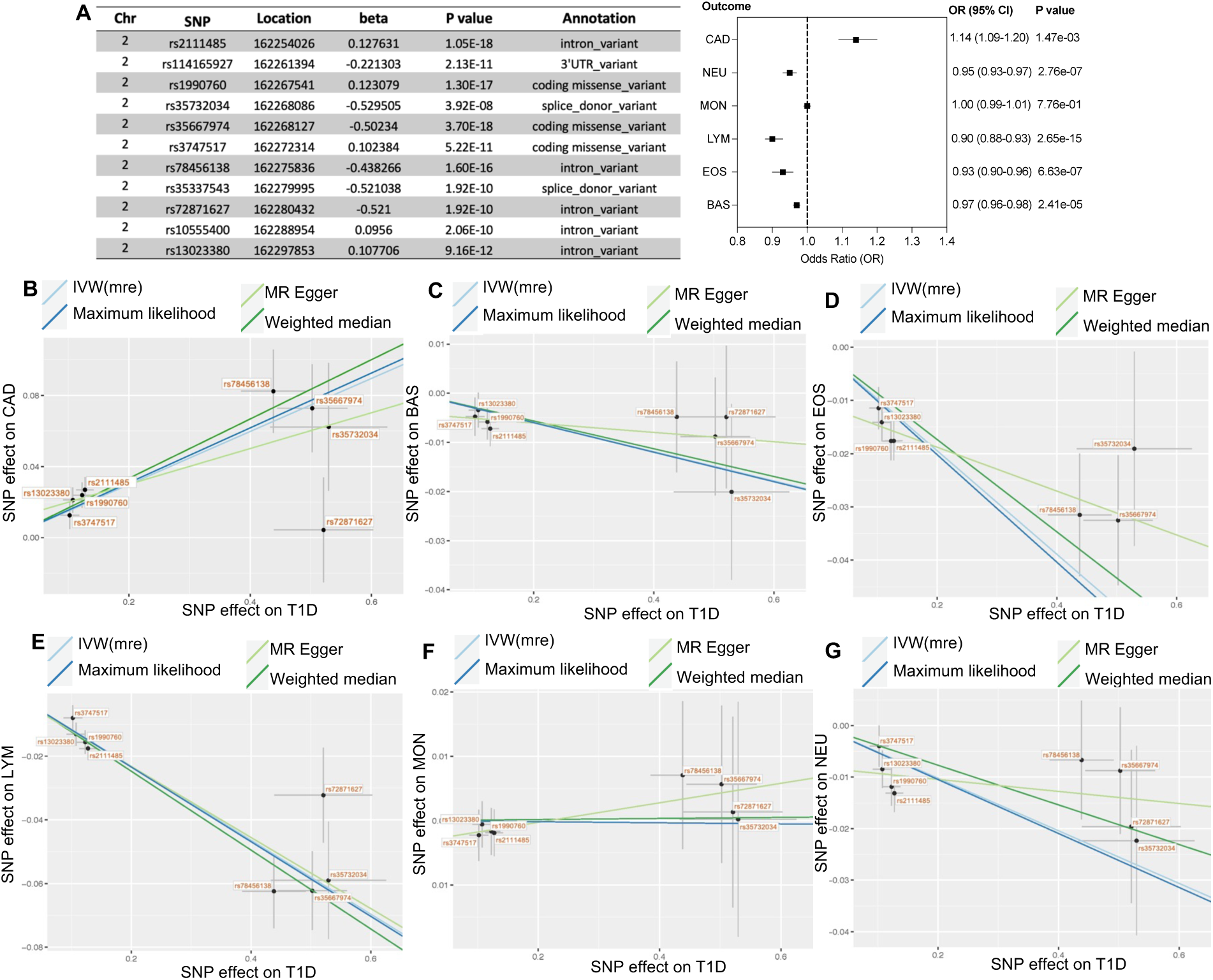
Association between T1D risk variants in IFIH1 and CAD and leukocyte counts. (**A**) Annotation of GWAS variants in *IFIH1* associated with T1D risk (p<5.0e-8) used as genetic instruments for MR analysis (left panel) and MR estimates of their effects on CAD and leukocyte counts (right panel). (**B-G)** Scatter plot showing the effect of T1D risk variation in *IFIH1* on **(B)** CAD, (**C**) basophil count, (**D**) eosinophil count, (**E**) lymphocyte count, and (**G**) neutrophil count.

### Genetically determined eosinophil, lymphocyte, and neutrophil counts is associated with T1D and CAD risk

The primary IVW-mre MR analysis showed that genetically predicted increased eosinophil count is associated with increased risk of T1D (OR = 1.67, CI = 1.52-1.85, P = 5.45E-25) and CAD (OR = 1.07; CI = 1.03-1.11, P = 2.02e-04; **Table 2 and Suppl Data 2**). These results were consistent in the MR-Egger test, except in the Weighted median test wherein eosinophil count did not associate significantly with CAD risk (OR = 1.02, CI = 0.96-1.09, P = 4.70e-01). In contrast to eosinophil count, genetic upregulation of the counts of basophils, neutrophils, and lymphocytes were inversely associated with T1D risk (OR_BAS_ = 0.70, CI = 0.58-0.83, P = 5.57e-05; OR_NEU_ = 0.82, CI = 0.75-0.90, P = 5.63e-05; OR_LYM_ = 0.67, CI = 0.61-0.73, P = 2.02e-19), while positively associating with CAD risk (OR_BAS_ = 1.23, CI = 1.13-1.35, P=2.47e-06; OR_NEU_ = 1.17, CI = 1.12-1.21, P= 5.02e-14; OR_LYM_ = 1.09, CI = 1.05-1.12, P = 2.67e-06; **Table 2 and Suppl Data 2**). Genetic predisposition to T1D was not associated with CAD and vice versa. The reverse MR analyses showed that genetic risk for T1D was associated with decreased lymphocyte count (OR = 0.98, CI = 0.98-0.99, P = 1.34e-08) and decreased neutrophil count (OR = 0.99, CI = 0.99-1.00, P = 1.38e-02), while increasing eosinophil count (OR = 1.01, CI = 1.01-1.02, P = 5.21e-05). Genetic predisposition to CAD did not have significant association with leukocyte counts.

T1D is considered a lymphocyte-mediated disease, therefore the apparently protective effect of increased lymphocyte counts was unexpected. To probe this finding further, we expanded our analyses to other related autoimmune diseases, including multiple sclerosis (MS) and rheumatoid arthritis (RA). Unlike T1D, genetic upregulation of lymphocyte count was associated with increased odds of MS (OR = 1.22, CI = 1.09-1.37, P = 7.61e-04) and no significant association with RA (OR = 0.97, CI = 0.85-1.11, P = 6.59e-01; **Suppl Data 2**).

## Discussion

This study demonstrates that T1D and CAD share a positive genetic architecture and that both diseases share significant genetic determinants with eosinophil and lymphocyte counts at the genome-wide level. Importantly, not only do T1D and CAD share genetic determinants with eosinophil and lymphocyte counts, but genetically determined counts of these leukocytes as well as neutrophils were also associated with both diseases in the same direction as the genetic correlations. At the locus-specific level, several loci were identified that had positive or negative genetic correlations between T1D and CAD, including loci that overlap *SH2B3*, *CTSH*, *MORF4L1*, *CFDP1*, *CTRB1*, *CTRB2*, and *IFIH1*. *Cis*-genetic regulation of the expression levels of these genes were also associated both diseases as well as leukocyte counts. GWAS had previously associated *CTSH*, *CTRB1*, *CTRB2*, and *IFIH1* with T1D [9, 11], and *CFDP1* and *MORF4L1* with CAD risk [23]. By analyzing summary level data, this study demonstrates that the loci that harbor these genes are shared between T1D and CAD and that genetic regulation of the expression levels of these is associated with both diseases. Overall, these findings suggest that shared genetic basis between T1D and CAD that affect leukocyte counts may link the two diseases. While genetically determined expression levels of *CTSH*, *MORF4L1*, and *CFDP1* had opposing associations with T1D and CAD, that for SH2B3, CTRB1, CTRB2, and IFIH1 had the same direction of effect on both diseases and could represent predictive markers or targets for both diseases. Deleterious coding variants in *CTSH* were associated with reduced risk of T1D [9], which is in line with the results in this study that genetic upregulation of *CTSH* expression levels in blood and pancreas increases T1D risk. This is also similar in the case of *SH2B3,* wherein GWAS *SH2B3* loss-of-function variants have been associated with increased T1D risk [9, 11] and atherosclerosis in murine models [35], while our analysis showed that genetic upregulation of *SH2B3* expression protects against T1D and CAD. Variants in *IFIH1* have been associated with many autoimmune diseases. In particular, loss-of-function variants have been associated with protection against T1D whereas gain-of-function variants increase T1D risk [11, 33]. Interestingly, not only did we find that cis-eQTL-mediated increased expression of *IFIH1* levels increase T1D and CAD risk, but also that GWAS variants in *IFIH1* that increased T1D risk also increased CAD risk while altering leukocyte counts. MR analysis that used *cis*-protein quantitative trait loci (*cis*-pQTL) as instruments associated genetic upregulation of chymotrypsinogen B1 (encoded by *CTRB1*) with reduced T1D risk [36]. Our studies using *cis*-eQTLs as instruments also showed that increased expression of *CTRB1* against T1D as well as CAD. Unlike *CTRB1*, genetic upregulation of *CTRB2* showed the opposite association with T1D and CAD, increasing the risk of both diseases.

Given that T1D is largely considered a lymphocyte-mediated disease, it was counterintuitive that T1D showed a negative genome-wide genetic correlation with lymphocyte count. Interestingly, genetically determined lymphocyte count had an inverse association with T1D risk, and vice versa, which is consistent the genetic correlations. T1D risk variants were shown to have a predominant enrichment in lymphoid enhancers, but it was unknown whether this relates to lymphocyte counts [10]. In addition to a genome-wide genetic correlation, several T1D risk loci, including *PTPN22*, *IL2RA*, *HORMAD2*, *KLRG1*/*CD69*, and *ZBTB9*/*TCP11* showed strong local genetic correlations with lymphocyte counts. Similar to lymphocyte counts, neutrophil count had an inverse association with T1D risk and displayed a negative genome-wide genetic correlation with T1D, albeit statistically insignificant. Clinical observations have documented reduced lymphocyte and neutrophil counts in patients with of T1D [16–19, 37]. Our findings thus suggest that these altered counts could contribute to disease.

Eosinophil count displayed significant genome-wide positive genetic correlation with both T1D and CAD and genetic upregulation of eosinophil count was positively associated with both diseases. Observational studies have associated eosinophil counts with CAD [38, 39] and shown that eosinophils from patients with T1D express high levels of myeloid alpha defensins and myelopoxidase, which could be pathogenic [40]. Thus, eosinophils may play a causative role in both T1D and CAD. In line with this proposition, we identified several T1D and/or CAD risk loci that showed significant local genetic correlations with eosinophil count, including *CLEC16A, IKZF4/SOUX, BLTP1/ADAD1, BACH2, ATXN2, and SMAD3*.

In summary, this study identifies shared genetic basis between T1D and CAD and implicate genetic regulation of leukocyte counts and gene expression levels as potential contributors to both diseases and their co-occurrence. A potential limitation to the findings in this study relates to potential confounders in MR analysis, such as genetic pleiotropy, which makes it problematic to conclude that the outcome is entirely due to the effect of the instruments on the exposure. Experimental perturbation of the shared genes between T1D and CAD in hematopoietic cells would also enable a better understanding of the genetic mechanisms that contribute to the co-occurrence of the two diseases through leukocytes.

## Data Availability

The source of publicly available data used for analyses in this study are described in the manuscript. All data produced in the present study are available upon reasonable request to the authors.

## Abbreviations

T1D: Type 1 diabetes
CAD: Coronary artery disease
GWAS: Genome-wide association studies
HSPCs: Hematopoietic stem and progenitor cells
LD: Linkage disequilibrium
LDSC: Linkage disequilibrium score regression
p-HESS: Heritability estimation from summary statistics
eQTL: Expression quantitative trait loci
BAS: Basophils
EOS: Eosinophils
LYM: Lymphocytes
Mon: Monocytes
NEU: Neutrophils
pQTL: Protein quantitative trait loci

## ACKNOWLEDGEMENTS

These studies were supported by JDRF (2-SRA-2022-1276-S-B).

## SUPPLEMENTARY TABLES AND FIGURES

**Suppl Table 1:** Genome-wide genetic correlations between leukocyte counts, T1D, and CAD.

| Trait 1 | Trait 2 | $r_g$ | $r_g$ SE | P value | $h^2$ | $h^2$ SE |
| --- | --- | --- | --- | --- | --- | --- |
| T1D | CAD | 0.0881 | 0.0335 | 8.60e-03** | 0.0604 | 0.0042 |
| T1D | BAS | 0.0153 | 0.0379 | 6.86E-01 | 0.0327 | 0.0035 |
| T1D | EOS | 0.0928 | 0.0346 | 7.20e-03** | 0.1289 | 0.0111 |
| T1D | LYM | -0.0523 | 0.0238 | 2.76e-02* | 0.1641 | 0.0127 |
| T1D | MON | -0.0099 | 0.0348 | 7.76E-01 | 0.1543 | 0.0143 |
| T1D | NEU | -0.0115 | 0.0207 | 5.80E-01 | 0.1191 | 0.0081 |
| CAD | BAS | 0.0858 | 0.0264 | 1.10e-03*** | 0.0324 | 0.0038 |
| CAD | EOS | 0.0923 | 0.0199 | 3.68e-06**** | 0.1275 | 0.0133 |
| CAD | LYM | 0.1761 | 0.0221 | 1.82e-15**** | 0.1629 | 0.0132 |
| CAD | MON | 0.1267 | 0.0205 | 6.35e-10**** | 0.1458 | 0.0132 |
| CAD | NEU | 0.1851 | 0.02 | 2.15e-20**** | 0.1145 | 0.0094 |

**Suppl Data 1: Local genetic *analyses by* ρ-HESS. (Sheet 1) *Genome-wide agnostic scan for significant local genetic correlations*.** Local heritability and genetic correlation estimates are shown for LD-independent blocks and pairs of traits with nominally significant local heritability and genetic correlation estimates. Estimates in red correspond to cases where the local genetic correlation p value exceeds a Bonferroni threshold for the number of LD-independent blocks in analysis (i.e. 0.05/1453). We recommend individually interpreting these Bonferroni-significant loci, and using the larger list for downstream bioinformatic analyses where a higher false discovery rate is tolerable. **(Sheet 2)**. ***Local genetic correlation analysis at preselected gene loci*.** Local heritability and genetic correlation estimate at each gene locus are shown for each pair of traits for the LD-independent block containing the gene, with corresponding p values (Bonferroni significance 0.05/36). Genetic correlation values are set to NA in cases where there is not nominally significant heritability for at least one of the two traits. SNPs with lowest p value per trait are also shown.

**Suppl Data 2.** Mendelian randomization estimates of association between cis-genetic regulation of the expression levels of shared genes between T1D and CAD on both diseases, and bi-directional MR estimates of the associations between genetically determined leukocyte counts and T1D and CAD.

## REFERENCES

1. Hippisley-Cox, J., C. Coupland, and P. Brindle, Development and validation of QRISK3 risk prediction algorithms to estimate future risk of cardiovascular disease: prospective cohort study. Bmj, 2017. 357: p. j2099.

2. Petrie, J.R. and N. Sattar, Excess Cardiovascular Risk in Type 1 Diabetes Mellitus. Circulation, 2019. 139(6): p. 744–747.

3. Bebu, I., et al., Genetic Risk Factors for CVD in Type 1 Diabetes: The DCCT/EDIC Study. Diabetes Care, 2021.

4. Rawshani, A., et al., Range of Risk Factor Levels: Control, Mortality, and Cardiovascular Outcomes in Type 1 Diabetes Mellitus. Circulation, 2017. 135(16): p. 1522–1531.

5. Eckel, R.H., K.E. Bornfeldt, and I.J. Goldberg, Cardiovascular disease in diabetes, beyond glucose. Cell Metab, 2021.

6. Astle, W.J., et al., The Allelic Landscape of Human Blood Cell Trait Variation and Links to Common Complex Disease. Cell, 2016. 167(5): p. 1415–1429.e19.

7. Jaiswal, S., et al., Clonal Hematopoiesis and Risk of Atherosclerotic Cardiovascular Disease. N Engl J Med, 2017. 377(2): p. 111–121.

8. Bick, A.G., et al., Inherited causes of clonal haematopoiesis in 97,691 whole genomes. Nature, 2020. 586(7831): p. 763–768.

9. Chiou, J., et al., Interpreting type 1 diabetes risk with genetics and single-cell epigenomics. Nature, 2021.

10. Onengut-Gumuscu, S., et al., Fine mapping of type 1 diabetes susceptibility loci and evidence for colocalization of causal variants with lymphoid gene enhancers. Nat Genet, 2015. 47(4): p. 381–6.

11. Robertson, C.C., et al., Fine-mapping, trans-ancestral and genomic analyses identify causal variants, cells, genes and drug targets for type 1 diabetes. Nature Genetics, 2021.

12. Örd, T., et al., Single-Cell Epigenomics and Functional Fine-Mapping of Atherosclerosis GWAS Loci. Circ Res, 2021. 129(2): p. 240–258.

13. Örd, T., et al., Dissecting the polygenic basis of atherosclerosis via disease-associated cell state signatures. The American Journal of Human Genetics.

14. Bao, E.L., et al., Inherited myeloproliferative neoplasm risk affects haematopoietic stem cells. Nature, 2020. 586(7831): p. 769–775.

15. Ulirsch, J.C., et al., Interrogation of human hematopoiesis at single-cell and single-variant resolution. Nat Genet, 2019. 51(4): p. 683–693.

16. Salami, F., et al., Complete blood counts with red blood cell determinants associate with reduced beta-cell function in seroconverted Swedish TEDDY children. Endocrinol Diabetes Metab, 2021. 4(3): p. e00251.

17. Salami, F., et al., Reduction in White Blood Cell, Neutrophil, and Red Blood Cell Counts Related to Sex, HLA, and Islet Autoantibodies in Swedish TEDDY Children at Increased Risk for Type 1 Diabetes. Diabetes, 2018. 67(11): p. 2329–2336.

18. Vecchio, F., et al., Abnormal neutrophil signature in the blood and pancreas of presymptomatic and symptomatic type 1 diabetes. JCI Insight, 2018. 3(18).

19. Valle, A., et al., Reduction of circulating neutrophils precedes and accompanies type 1 diabetes. Diabetes, 2013. 62(6): p. 2072–7.

20. Madjid, M., et al., Leukocyte count and coronary heart disease: implications for risk assessment. J Am Coll Cardiol, 2004. 44(10): p. 1945–56.

21. Summers, C., et al., Neutrophil kinetics in health and disease. Trends Immunol, 2010. 31(8): p. 318–24.

22. Kachuri, L., et al., Genetic determinants of blood-cell traits influence susceptibility to childhood acute lymphoblastic leukemia. Am J Hum Genet, 2021. 108(10): p. 1823–1835.

23. van der Harst, P. and N. Verweij, Identification of 64 Novel Genetic Loci Provides an Expanded View on the Genetic Architecture of Coronary Artery Disease. Circ Res, 2018. 122(3): p. 433–443.

24. Bulik-Sullivan, B., et al., An atlas of genetic correlations across human diseases and traits. Nat Genet, 2015. 47(11): p. 1236–41.

25. Shi, H., et al., Local Genetic Correlation Gives Insights into the Shared Genetic Architecture of Complex Traits. Am J Hum Genet, 2017. 101(5): p. 737–751.

26. Berisa, T. and J.K. Pickrell, Approximately independent linkage disequilibrium blocks in human populations. Bioinformatics, 2016. 32(2): p. 283–5.

27. LeBlanc, M., et al., Identifying Novel Gene Variants in Coronary Artery Disease and Shared Genes With Several Cardiovascular Risk Factors. Circ Res, 2016. 118(1): p. 83–94.

28. Patrick, M.T., et al., Shared genetic risk factors and causal association between psoriasis and coronary artery disease. Nat Commun, 2022. 13(1): p. 6565.

29. Maurano, M.T., et al., Systematic localization of common disease-associated variation in regulatory DNA. Science, 2012. 337(6099): p. 1190–5.

30. Kim-Hellmuth, S., et al., Cell type-specific genetic regulation of gene expression across human tissues. Science, 2020. 369(6509).

31. Bowden, J., et al., Improving the visualization, interpretation and analysis of two-sample summary data Mendelian randomization via the Radial plot and Radial regression. Int J Epidemiol, 2018. 47(6): p. 2100.

32. Bowden, J., et al., Consistent Estimation in Mendelian Randomization with Some Invalid Instruments Using a Weighted Median Estimator. Genet Epidemiol, 2016. 40(4): p. 304–14.

33. Nejentsev, S., et al., Rare variants of IFIH1, a gene implicated in antiviral responses, protect against type 1 diabetes. Science, 2009. 324(5925): p. 387–9.

34. Liu, S., et al., IFIH1 polymorphisms are significantly associated with type 1 diabetes and IFIH1 gene expression in peripheral blood mononuclear cells. Hum Mol Genet, 2009. 18(2): p. 358–65.

35. Wang, W., et al., LNK/SH2B3 Loss of Function Promotes Atherosclerosis and Thrombosis. Circ Res, 2016. 119(6): p. e91–e103.

36. Yazdanpanah, N., et al., Clinically Relevant Circulating Protein Biomarkers for Type 1 Diabetes: Evidence From a Two-Sample Mendelian Randomization Study. Diabetes Care, 2022. 45(1): p. 169–177.

37. Harsunen, M.H., et al., Reduced blood leukocyte and neutrophil numbers in the pathogenesis of type 1 diabetes. Horm Metab Res, 2013. 45(6): p. 467–70.

38. Gao, S., et al., Eosinophils count in peripheral circulation is associated with coronary artery disease. Atherosclerosis, 2019. 286: p. 128–134.

39. Tanaka, M., et al., Eosinophil count is positively correlated with coronary artery calcification. Hypertens Res, 2012. 35(3): p. 325–8.

40. Neuwirth, A., et al., Eosinophils from patients with type 1 diabetes mellitus express high level of myeloid alpha-defensins and myeloperoxidase. Cell Immunol, 2012. 273(2): p. 158–63.

